# Predictors of Delayed or Absent Measles, Mumps, and Rubella Vaccination in the US, 2018–2025

**DOI:** 10.1101/2025.06.09.25329274

**Authors:** Nina B. Masters, Brianna M. Goodwin Cartwright, Patricia J. Rodriguez, Karen M. Gilbert, Duy Do, Nicholas L. Stucky

## Abstract

**Importance:** 1,168 measles cases have been reported as of June 6, 2025, and cases for the year are likely to soon exceed the highest levels seen since elimination in 2000. MMR (measles, mumps, rubella) vaccination rates have declined, and predictors of delayed and missed vaccination since the COVID-19 pandemic are not well explored.

**Objective:** Characterize rates and trends of timely MMR vaccination among children and assess predictors of late and non-vaccination by two years of age.

**Design:** In this retrospective cohort study, infants who accessed routine care within the first two months, first year, and second year of life were followed for 24 months to assess vaccination outcomes between January 1, 2018, and April 30, 2025.

**Setting:** Children seeking care within a healthcare system partnering with Truveta, an electronic health record database from a collective of US health care systems.

**Participants:** This study included 321,743 children under the age of two years.

**Exposures:** The primary exposure(s) were timely receipt of routine 2- and 4-month immunizations and adherence to the AAP well child visit schedule.

**Main Outcome(s) and Measure(s):** The primary outcome was timely, late, or no receipt of MMR by two years of age. Associations with primary exposures and sociodemographic factors were modeled using mixed effect logistic regression with state-level random effects. Models were stratified by pre-versus post-COVID-19 MMR eligibility, with results after the COVID-19 pandemic reported as primary.

**Results:** In this study of 321,743 children, 78.4% received their first MMR on time, rising from 75.6% (12,840/16,978) in 2018, to 79.9% (39,739/49,767) in 2021, then declining to 76.9% (40,306/52,388) in 2024. The strongest predictor of no MMR vaccination by two years was late administration of a child’s second-month vaccines (aOR 6.96 [6.60-7.34]) and fourth-month vaccines (aOR 6.16 [5.84,6.50]).

**Conclusions and Relevance:** In this cohort study, most children received their MMR vaccine on time, but the proportion of children not receiving MMR by two years of age has steadily grown each year since the COVID-19 pandemic. Children who did not receive their 2- and 4-month vaccines on time were significantly more likely to not receive any MMR by two years, highlighting early opportunities for intervention.

## Introduction

Vaccination is one of the greatest accomplishments of public health, resulting in dramatic declines in childhood morbidity and mortality.^1–3^ However, vaccine hesitancy has been increasing both globally^4,5^ and in the United States.^6–8^ The COVID-19 pandemic may have had a ‘spillover effect’, with COVID-19-related vaccine hesitancy causing increased hesitancy towards other vaccines, including routine immunizations.^9^ Parents with vaccine hesitancy are more likely to intentionally delay vaccinations for their children, prolonging their susceptibility to infectious, vaccine-preventable diseases.^10^ Measles, declared eliminated from the US in 2000, has resurged in 2025,^11^ along with pertussis,^12^ a signal that diseases controlled by effective vaccination campaigns with high population coverage appear to again be spreading more widely. Given that 2025 has seen the first deaths from measles in the US in a decade,^11^ it is essential to understand rates and trends of measles, mumps, and rubella (MMR) vaccination among children, and explore predictors of on time, late, or non-vaccination.

Children receive several lifesaving vaccines in the first two years of life,^13^ including Hepatitis B vaccination at birth, Rotavirus, Diphtheria Tetanus, and acellular Pertussis (DTaP), *Haemophilus influenzae* type b (*Hib*), Pneumococcal Conjugate Vaccine (PCV), and inactivated poliovirus vaccines (IPV), and the measles-mumps-rubella (MMR) and varicella vaccines. Vaccines are recommended to be administered according to a standard scheduled from the CDC.^13^ Alternative vaccine schedules, such as the one popularized by Dr. Bob Sears, advocate for more spaced-out intervals for immunization: delaying polio vaccination until 9 months and Hepatitis B vaccination until over 2 years of age. These delayed schedules lead to prolonged susceptibility among young children most vulnerable to severe disease.^14^ Recently published research signaled better adherence to the recommended vaccine schedule just prior to the COVID-19 pandemic, with fewer children following alternative schedules^15^ and a greater proportion of children receiving recommended vaccines by 19 months.^16^ However, studies using data since the COVID-19 pandemic are needed, as timely adherence to childhood vaccinations may have changed.

Understanding predictors of untimely MMR vaccination is important to aid pediatricians in having early conversations with parents about vaccinations, helping to allay hesitancy and ensure children are on track. Prior studies using electronic health record (EHR) data have shown that children who were not up to date on early immunizations^17,18^ were more likely to be delayed on the rest of their vaccines. However, these studies have not explored predictors of untimely immunization since the COVID-19 pandemic – a period marked by potentially increased vaccine hesitancy and altered healthcare access. We explored rates and predictors of timely and delayed childhood immunization among children under 2 years.

## Methods

### Data

The study used a subset of Truveta Data, an EHR database with over 120 million individuals from a collective of healthcare systems in the United States.^19^ The data provided structured demographics, encounter details, diagnoses, immunizations, and procedures relevant to this analysis. Data undergo syntactic and semantic normalization and are then de-identified by expert determination under the HIPAA Privacy Rule. Data were accessed May 16, 2025. This analysis excluded states where healthcare systems did not provide encounter data for the month preceding data access, or did not provide procedure data. This study used de-identified patient records and therefore did not require Institutional Review Board approval.

### Population and Design

The study included 321,743 children born from 2017-2023 who received regular care within a Truveta constituent healthcare system, defined by outpatient visits at 0-2 months (0-90 days), 3-11 months (91-364 days), 12-21 months (365-668 days), and 22-25 months (669-790 days) of age. These four visits in the first two years of life were selected to exclude patients not receiving regular care from Truveta’s healthcare systems, and to capture two full years of follow-up after birth. For children with more than one visit in each window, the last visit was used. Individuals with missing sex (n = 10) were removed from analysis.

### Vaccinations

Vaccines in the childhood primary series, including *Hib*, DTaP, PCV, IPV, and Rotavirus vaccines, in addition to the primary outcome, MMR, were assessed based on vaccine administered (CVX) and Current Procedural Terminology (CPT) codesets (Supplementary Table S1). Vaccines received between birth and two years of age (730 days) were included. Vaccines were classified as appropriately administered, and bucketed into early, on-time, or late given CDC recommended age and dosing intervals, following ACIP guidelines with a 4-day grace period prior to the minimum age of administration.^13^ Vaccinations received prior to the earliest minimum age of administration (i.e. MMR vaccines received before 182 days) or vaccinations received in violation of CDC minimum dosage intervals^20^ were removed due to potential data errors.

### Statistical Analysis

Time series plots and monthly rate calculations of vaccinations by timeliness category were calculated using a denominator of children who met the eligibility criteria for each outpatient visit. MMR vaccinations were plotted using the year of the child’s last visit in the 12–21-month period, while 2-and 4-month vaccinations were plotted using the child’s birth month shifted by 2- and 4-months, respectively. A chi-square test was used to assess the significance of these time trends.

Descriptive analyses and multilevel logistic regression models with state-level random effects were used to evaluate predictors of delayed MMR or non-vaccination, compared to timely vaccination or any vaccination, respectively. Statistical analyses adjusted for potentially confounding variables such as sex, race, ethnicity, state of residence (used as a random effect to control for state level clustering of outcomes), an indicator for rural/urban residence based on Federal Office of Rural Health Policy Data files from 2024,^21^ and a utilization-based measure of whether or not children adhered to the American Academy of Pediatrics (AAP) schedule of well child visits in the first two years of life:^22^ a visit during the first 3-5 days, at one month, two months, four months, six months, nine months, 12 months, 15 months, 18 months, and 24 months. A 7-day grace period was applied to the start and a 30-day grace period to the end of these date windows. The 3–5-day visit was omitted as children needing NICU or specialized care would be unexpected to receive an outpatient visit during this window.

Models were stratified by the timing of the COVID-19 pandemic: if a child’s 12–21-month visit occurred prior to March 2020, the visit was ‘pre-pandemic’ and visits after September 2020 were ‘post-pandemic’. Visits during March – August 2020 were excluded from both time periods due to pandemic interruptions, with substantial declines in well child visits in the spring and summer of 2020.^23^ For brevity, model results after the pandemic are included in text, but all odds ratios and confidence intervals are shown in Figures and Supplementary Tables. Analyses were conducted in R version 4.4.1 within a cloud-based notebook environment leveraging Apache Spark.

## Results

### Predictors of MMR Timeliness

In this population of 321,743 children under two years of age, 48.4% were female and 51.6% male, and 60.5% of the population was White, 10.1% Black or African American, 6.2% Asian, 8.3% other race, and 14.8% unknown race. Overall, 77.7% of children in the study resided in urban areas, 19.0% in rural areas, and 3.3% in an area of unknown urbanicity; and 48.9% of children adhered to the AAP’s recommended well-child visit schedule in the first two years of life. Overall, 1.0% of children received MMR early, 78.4% of children received their MMR vaccine on-time, 13.9% received MMR late, and 6.7% did not have an MMR recorded by 2 years.

### Trends of MMR Vaccination by 2 Years of Age

Timely MMR coverage rates changed significantly over time (p-value <0.001), rising from 75.6% (12,840/16,978) in 2018, to 79.9% (39,739/49,767) in 2021, then declining to 76.9% (40,306/52,388) in 2024. (Figure 1, Table S2). The post-2021 decline in timely MMR vaccination corresponded to an increase in the proportion of children with no MMR vaccine recorded by 2 years, which increased from 5.3% (2,617/49,760) in 2020 to 7.7% (4,044/52,388) in 2024. Late MMR vaccination stayed relatively constant at around 14% over the study period, and early (6-11 month) vaccinations remained rare (<1%), aside from an increase to 2.9% for children with a 12-21 month visit in 2020.

**Fig 1.**
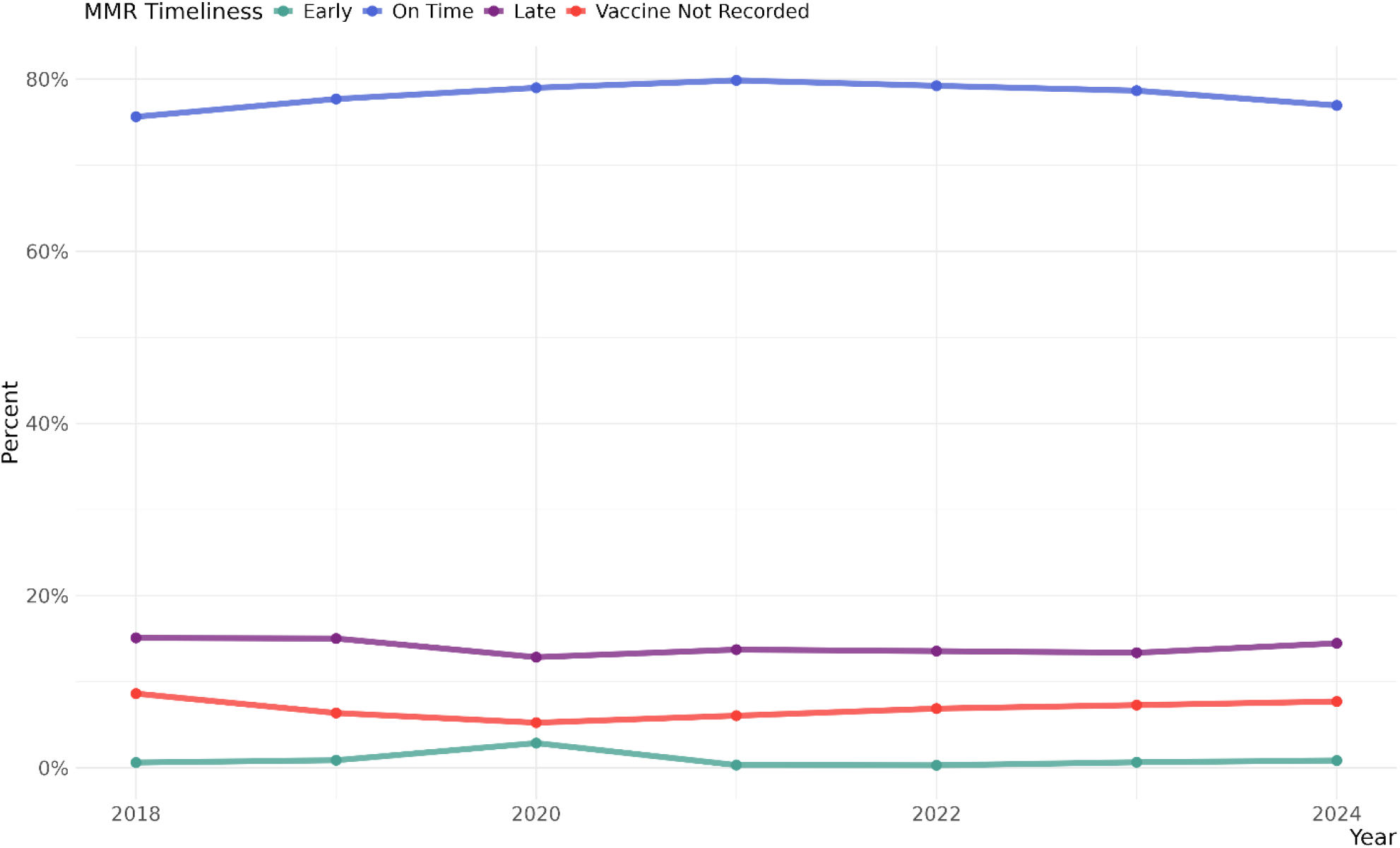
Proportion of Children Receiving their First MMR Vaccine Early, On-Time, Late, or Not at all by 2 Years of Age by the Year of their 12–21-month visit.

**Table 1.** Sociodemographic, Health Care Utilization, and First MMR Timeliness by Timing of 12-21 Month Visit.

|  | Timing of 12-21 Month Outpatient Visit* |  |  |  |
| --- | --- | --- | --- | --- |
|  | Overall | Feb 2020 and Before | Mar - Aug 2020 | Sep 2020 and Later |
|  | N = 321,743 | N = 74,177 | N = 22,866 | N = 224,700 |
| **Gender** |  |  |  |  |
| Female | 155,726 (48.4%) | 36,205 (48.8%) | 11,207 (49.0%) | 108,314 (48.2%) |
| Male | 166,017 (51.6%) | 37,972 (51.2%) | 11,659 (51.0%) | 116,386 (51.8%) |
| **Race** |  |  |  |  |
| Asian | 19,832 (6.2%) | 5,113 (6.9%) | 1,523 (6.7%) | 13,196 (5.9%) |
| Black or African American | 32,656 (10.1%) | 6,878 (9.3%) | 2,059 (9.0%) | 23,719 (10.6%) |
| White | 194,762 (60.5%) | 46,860 (63.2%) | 14,468 (63.3%) | 133,434 (59.4%) |
| Other | 26,755 (8.3%) | 6,733 (9.1%) | 1,968 (8.6%) | 18,054 (8.0%) |
| Unknown | 47,738 (14.8%) | 8,593 (11.6%) | 2,848 (12.5%) | 36,297 (16.2%) |
| **Ethnicity** |  |  |  |  |
| Hispanic or Latino | 42,603 (13.2%) | 8,359 (11.3%) | 2,881 (12.6%) | 31,363 (14.0%) |
| Not Hispanic or Latino | 222,169 (69.1%) | 54,499 (73.5%) | 16,362 (71.6%) | 151,308 (67.3%) |
| Other | 5,119 (1.6%) | 973 (1.3%) | 323 (1.4%) | 3,823 (1.7%) |
| Unknown | 51,852 (16.1%) | 10,346 (13.9%) | 3,300 (14.4%) | 38,206 (17.0%) |
| **Rural or Urban Residence** |  |  |  |  |
| Rural | 61,074 (19.0%) | 15,215 (20.5%) | 4,349 (19.0%) | 41,510 (18.5%) |
| Urban | 249,991 (77.7%) | 56,943 (76.8%) | 17,842 (78.0%) | 175,206 (78.0%) |
| Unknown | 10,678 (3.3%) | 2,019 (2.7%) | 675 (3.0%) | 7,984 (3.6%) |
| **AAP Visit Schedule** |  |  |  |  |
| AAP Schedule Followed | 157,427 (48.9%) | 36,889 (49.7%) | 11,033 (48.3%) | 109,505 (48.7%) |
| Not Followed | 164,316 (51.1%) | 37,288 (50.3%) | 11,833 (51.7%) | 115,195 (51.3%) |
| **First MMR Vaccine Timing** |  |  |  |  |
| Early: 6-11 months | 3,106 (1.0%) | 828 (1.1%) | 829 (3.6%) | 1,449 (0.6%) |
| On-Time: 12-15 months | 252,250 (78.4%) | 57,502 (77.5%) | 18,109 (79.2%) | 176,639 (78.6%) |
| Late: 16-23 months | 44,718 (13.9%) | 10,798 (14.6%) | 2,787 (12.2%) | 31,133 (13.9%) |
| No Vaccine Recorded by 2 Years | 21,669 (6.7%) | 5,049 (6.8%) | 1,141 (5.0%) | 15,479 (6.9%) |

\*Last visit in 12–21-month window if more than one visit occurred

### Rates of Early Childhood Vaccinations

Time trends in the percent of children who received routine early childhood immunizations, such as IPV, DTaP, Hib, PCV, and Rotavirus vaccines (administered at 2 and 4 months) are shown in Figure 2.

**Fig 2.**
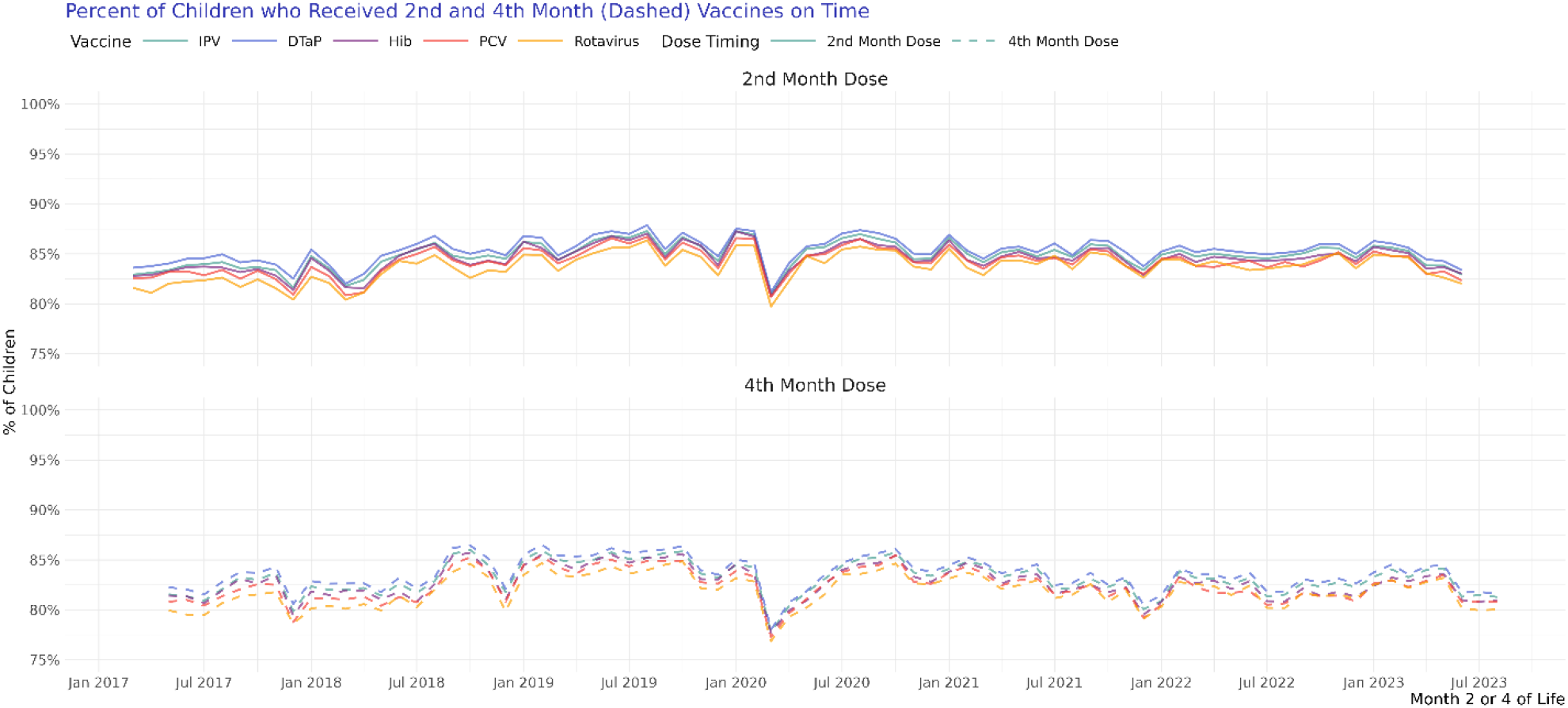
Proportion of children who received their 2^nd^ and 4^th^ month vaccines by the recommended age, with the X-axis representing birth month shifted forward by 2 and 4 months, respectively, to allow for visualization of vaccination in calendar time.

Overall, timely completion of 2^nd^ month vaccines (85.3% for DTaP, 84.9% for Polio, 84.6% for Hib, 84.2% for PCV and 83.7% for Rotavirus) was similar to 4^th^ month vaccines (83.6% for DTaP, 83.1% for Polio, 82.7% for Hib, 82.3% for PCV, and 81.9% for Rotavirus). Timely receipt of both 2^nd^ and 4^th^ month vaccines declined after the COVID-19 pandemic (Table S3, S4). Early vaccination represented approximately 6.3% of doses for all 2^nd^ month vaccines and 2.2% of doses for all 4^th^ month vaccines.

Timely first DTaP vaccinations declined from 86.5% in 2019 to 84.3% in 2023, while timely second DTaP vaccinations declined from 84.5% in 2020 to 82.2% in 2023 (Table S3, S4). Overall, on-time DTaP rates were highest and Rotavirus were lowest, and these trends were consistent over time.

### Predictors of No or Untimely MMR by 2 Years of Age

After the COVID-19 pandemic, the strongest predictors of not receiving any MMR vaccination compared to any receipt, were late 2^nd^ (aOR 6.96 [95% CI: 6.60-7.34]) and 4^th^ month vaccines (aOR 6.16 [95% CI: 5.84-6.50]) (Figure 3, Table S5). From 2022 to 2024, children had an increased odds of non-vaccination, with children eligible in 2024 having 1.35 [95% CI: 1.23-1.47] times the odds of non-vaccination by two years compared to children in 2020, adjusted for all other factors. Male children were slightly more likely to be unvaccinated (aOR 1.12 [95% CI: 1.07-1.16]), as were children living in rural areas (aOR 1.09 [95% CI: 1.03-1.15]), areas of unknown urbanicity (aOR 1.45 [95% CI: 1.18-1.79), White children (aOR 1.48 [95% CI: 1.39-1.59]), and children who were non-Hispanic or Latino (aOR 1.51 [95% CI: 1.41-1.61]) or unknown ethnicity (aOR 1.43 [95% CI: 1.31, 1.57]). Children who completed the AAP routine visit schedule had lower odds of non-vaccination (aOR 0.37 [95% CI: 0.36-0.39]).

**Fig 3.**
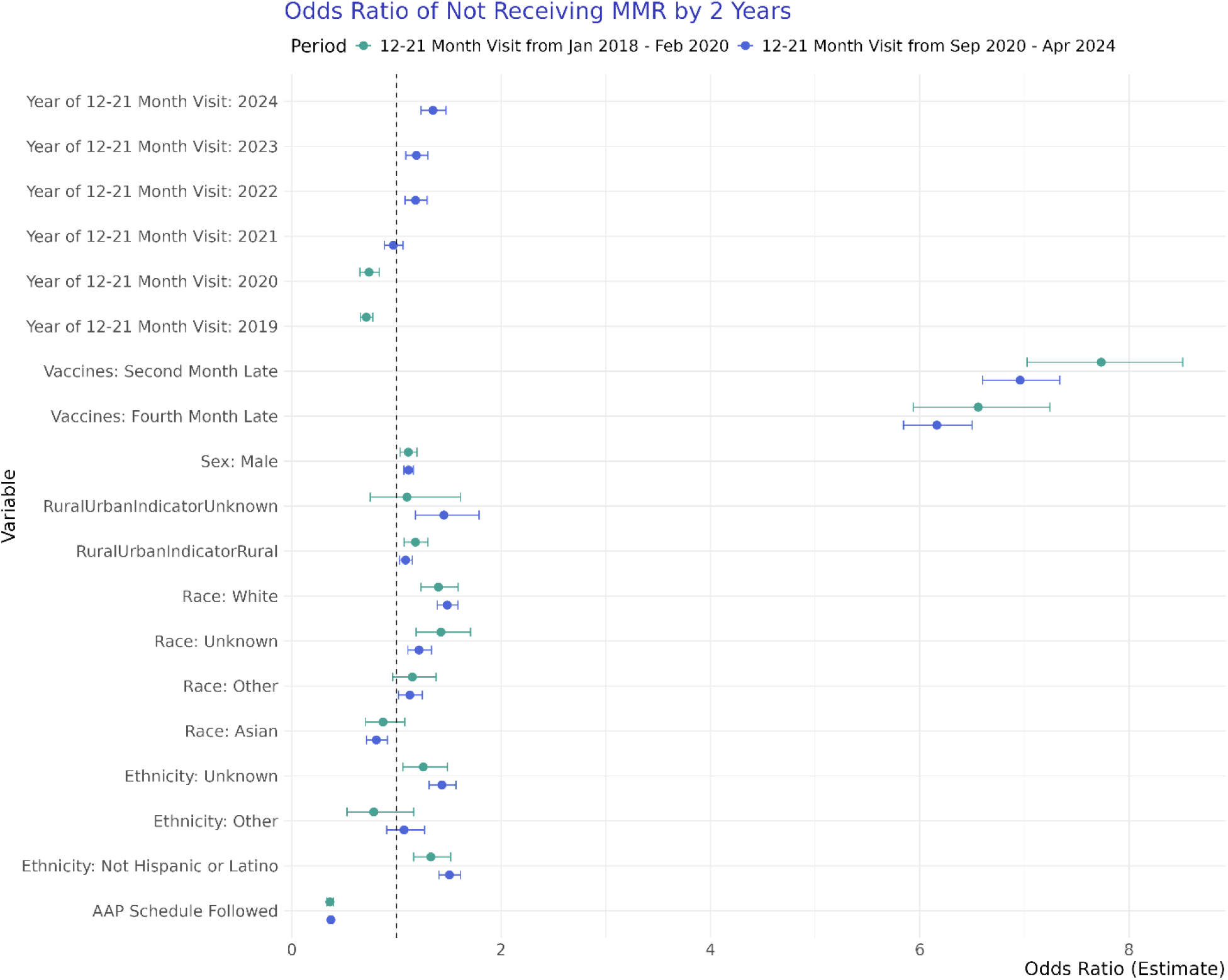
Odds Ratios of Not Receiving any MMR by Two Years of Age vs. Receiving Any MMR, Stratified by 12-21-month Visit Timing Before or After the COVID-19 Pandemic.

Analyses of late vs. on-time vaccination included 68,300 individuals before the pandemic and 207,772 after the pandemic, and late 4^th^ month vaccine receipt was the single strongest predictor (aOR 2.55 [95% CI: 2.43-2.67]), followed by late 2^nd^ month vaccines (aOR 1.47 [95% CI: 1.37-1.57]) (Figure 4, Table S6). In this analysis, children in rural settings (aOR 0.69 [95% CI: 0.66-0.71]), Asian children (aOR 0.47 [95% CI: 0.43-0.51]), and children who adhered to the AAP visit schedule (aOR 0.61 [95% CI: 0.59-0.63]) in the first two years of life were substantially more likely to receive timely MMR.

**Fig 4.**
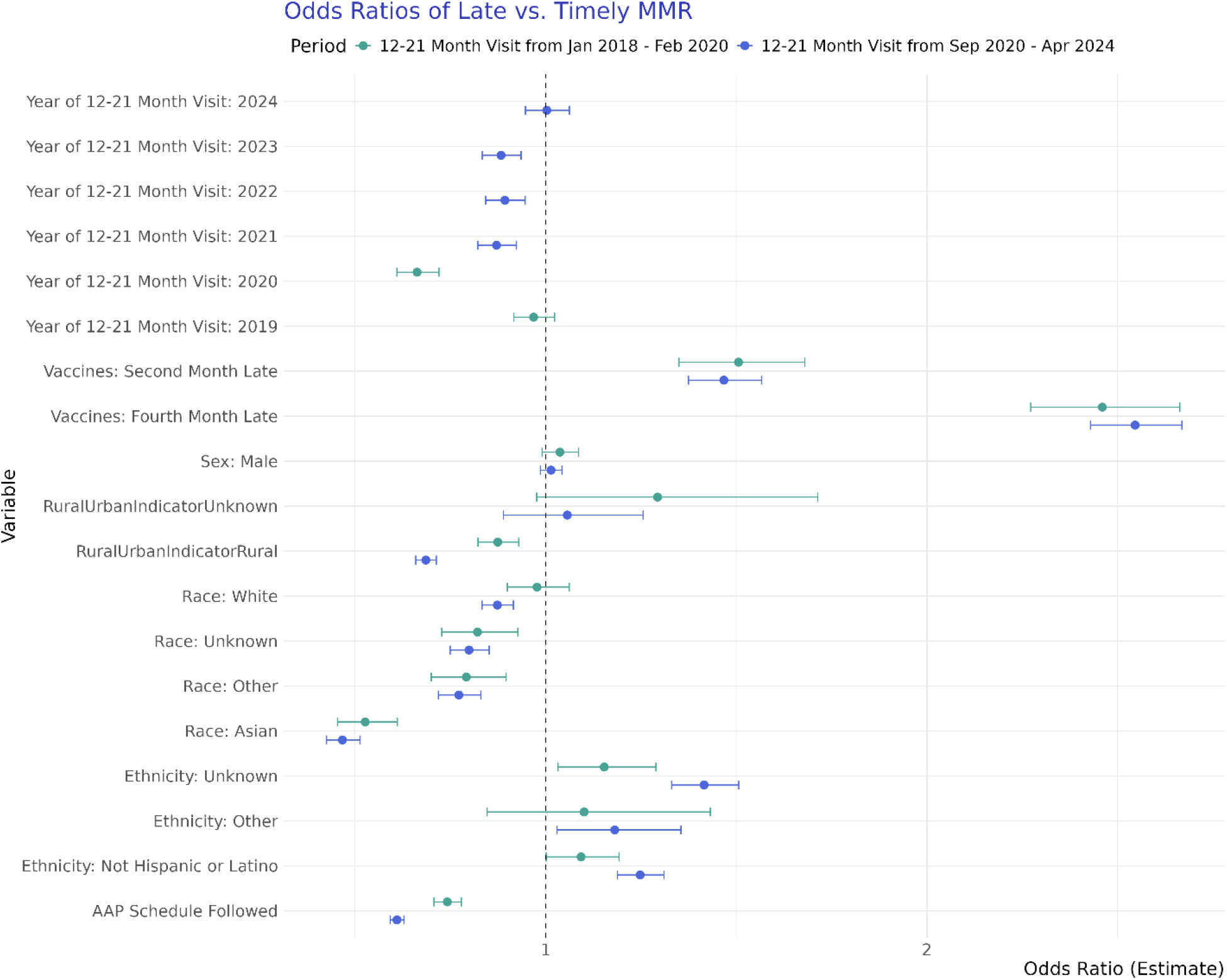
Odds Ratios of Late MMR Vaccination (Received at 16-23 months) Compared to On-Time MMR Vaccination, Stratified by 12-21-month Visit Timing Before or After the COVID-19 Pandemic.

## Discussion

In this large, EHR-based study of over 320,000 children, routine childhood vaccination rates and vaccine timeliness have been declining since 2021 for all vaccines assessed, indicating increased population-level susceptibility to vaccine-preventable diseases. Timely first-dose MMR vaccination rates among children decreased by three percentage points from 2021 to 2024, a worrisome decline, especially amid increase measles outbreaks in the US in 2025.^24^ This trend mirrors that reported by CDC^25^ where MMR coverage among children under two years declined from 92% for children born in 2018-19 to 90.3% for children born in 2020-21, but shows that this trend is continuing for children born in 2023.^26^ Our results also complement a recent analysis using county-level kindergarten data that showed continued declines in coverage through the 2024 school year for 2-doses of MMR.^27^ This population-level decline in timely vaccination may have contributed to rising pockets of susceptibility across the US, allowing for measles to spread more effectively.

Both before and since the COVID-19 pandemic, our results show that early vaccination decisions in a pediatric journey, specifically whether a child received their 2- and 4-month routine vaccines on-time (Hib, DTaP, Polio, Rotavirus, and PCV), are a strong predictor of MMR vaccination outcomes. Since the pandemic, children who received their second month on time had nearly seven times higher odds of receiving the MMR vaccine by age two, and two and a half times higher odds of receiving the MMR vaccine on time compared to those who did not. This highlights very early opportunities for intervention to address vaccine delay and potentially get children back on track. Research has shown that provider conversation style with parents has a strong impact on vaccine uptake at a pediatric visit.^28,29^ Specifically, the use of presumptive conversation styles around vaccines, i.e. “your child is getting two shots today” leads to much higher vaccine uptake (over 17-fold higher) than participatory conversations, i.e. “do you want to get vaccines for your child today?”.^30^ Additional integrated solutions within the EHR itself may also help, such as adding flags to children’s charts when they have missed milestones for immunizations, indicating the child is at risk to fall further behind the recommended schedule.

Our findings that male children had lower odds of receiving an MMR by two years of age may point to vaccine hesitancy regarding the debunked, fraudulent 1998 paper linking MMR to autism spectrum disorder (ASD).^31–33^ ASD occurs more frequently in male children,^34^ and thus parents may delay vaccination if they are very concerned about ASD, as a 2023 systematic review found that fear of autism was the most cited reason for hesitancy about the MMR vaccine.^35^ While studies have reported mixed findings on the likelihood of non-vaccination by race/ethnicity,^35,36^ our study found that White and non-Hispanic or Latino children were more likely to be unvaccinated for MMR, pointing to increased hesitancy among these groups in a care-seeking population. Interestingly, our analysis of late vs. timely MMR vaccination showed opposite associations from the non-vaccination analysis, with white race and rural residence both associated with higher odds of timely vaccination. This may point to fundamental differences between populations who delay vaccinations but ultimately vaccinate their children by two years of age (i.e. due to child sickness) vs. those who do not vaccinate their children by two years.

Additionally, because we required four outpatient visits for children in the first two years of life, our results may reflect populations without significant access challenges, whose decisions are motivated by vaccine hesitancy.

Declining timeliness in other vaccines received early in childhood (at 2- and 4-months of age) is also a concerning finding, especially amid increased transmission and infant deaths from pertussis reported in 2025.^37,38^ Rising pertussis incidence signals that vaccine-preventable diseases in general, not just measles, are now spreading more widely in the US due to decreased population immunity. Because the inclusion criteria for this study required four outpatient medical visits across a child’s first two years of life, we expect rates of timely and overall vaccination to be higher in this study than in the overall US population, as the study population represents those who seek and access care. By including individuals with consistent medical care over their first two years of life, the results of this analysis likely highlight decreases in timely vaccination attributed to rising vaccine hesitancy more than access issues.

Vaccine hesitancy is a complex issue, with multifactorial influences, including personal beliefs, access to health care, and additional sources of hesitancy and delays due to the COVID-19 pandemic.^6,36^ The majority of measles cases and hospitalizations in the US in recent years have occurred in children, notably in children under five years of age.^11,39^ Measles vaccination, provided through the MMR vaccine, is required for kindergarten entry in all states in the US, and thus the majority of under-immunization is likely to be concentrated in children under five years who have not yet started school. Young children are also most likely to experience severe disease and complications from measles,^40,41^ making under-immunization in this age group particularly concerning, and highlighting the importance of identifying potential intervention opportunities.

### Strengths and Limitations

This study has several strengths. First, the analysis utilized a large and recent cohort of children with MMR vaccinations administered through April 2025, providing extremely recent insight into a large, geographically diverse population of children. By incorporating state of residence as a random effect, clustering of vaccination outcomes, including variability in state laws regarding vaccine exemptions, was accounted for in analyses. This study allowed for an assessment of sociodemographic, geographic, and early vaccination events with two-year continuous follow-up, allowing for individual patient trajectories and vaccination decisions to be incorporated into analyses and minimizing the risk of bias from loss-to-follow-up. Finally, this study was able to incorporate vaccinations recorded both through immunization codes (CVX codes) and procedure data (CPT codes), as different health systems may rely on different modalities to capture immunizations in a patient record.

Our study is also subject to several limitations. Because the population was followed up for two years with repeat healthcare interactions, this is likely not representative of all children in the US; this analysis reflects a population with care through the first two years of life. As a result, this population may have higher care-seeking and higher vaccination coverage than the general population, however we allowed for flexibility in the timing of visit inclusion criteria to try to capture as many children as possible. We expect children without a medical home or adequate access to care may have experienced an even more significant drop in vaccination coverage since the pandemic than the results we see in our cohort. While most childhood vaccinations are administered in the healthcare system, vaccinations received by children at another provider outside of a Truveta constituent health facility, during travel, or at a pharmacy would not be captured in this analysis. Assuming the rate of vaccination outside the healthcare system was static over time, this may have impacted overall vaccination rates, but not trends. The study used clinical EHR data, which are collected during routine clinical care visits, not for the purposes of research.

However, we expect misclassification of administered childhood immunizations to be very uncommon, as these are procedures/recorded vaccinations administered in the clinical setting.

## Conclusions

The US has entered an increased landscape of surveillance and data uncertainty, as well as a moment of vaccine misinformation proliferation. This study shows that childhood vaccine rates and timeliness have been declining since the COVID-19 pandemic, and that staying on schedule early is most predictive of receiving a timely MMR vaccine, the best protection against the measles outbreaks that are spreading rapidly across the US. This study also highlights the importance of using electronic health record data both for timely vaccine surveillance, to identify opportunities for intervention, and to provide better care to patients.

## Data Availability

The data used in this study are available to all Truveta subscribers and may be accessed at studio.truveta.com.

## Acknowledgements

The authors thank Sam Gratzl for providing code review and optimization within the Apache Spark platform.

## References

1. Zhou, F. Health and Economic Benefits of Routine Childhood Immunizations in the Era of the Vaccines for Children Program — United States, 1994–2023. MMWR Morb. Mortal. Wkly. Rep. 73, (2024).

2. Roush, S. W., Murphy, T. V. & Vaccine-Preventable Disease Table Working Group, and the. Historical Comparisons of Morbidity and Mortality for Vaccine-Preventable Diseases in the United States. JAMA 298, 2155–2163 (2007).

3. Orenstein, W. A. & Ahmed, R. Simply put: Vaccination saves lives. Proc. Natl. Acad. Sci. U. S. A. 114, 4031–4033 (2017).

4. Dubé, E. et al. Vaccine hesitancy. Hum. Vaccines Immunother. 9, 1763–1773 (2013).

5. World Health Organization. Vaccine hesitancy: A growing challenge for immunization programmes. https://www.who.int/news/item/18-08-2015-vaccine-hesitancy-a-growing-challenge-for-immunization-programmes.

6. Larson, H. J., Gakidou, E. & Murray, C. J. L. The Vaccine-Hesitant Moment. N. Engl. J. Med. 387, 58–65 (2022).

7. Kaushik, A. et al. Pediatric Vaccine Hesitancy in the United States—The Growing Problem and Strategies for Management Including Motivational Interviewing. Vaccines 13, 115 (2025).

8. Cunningham, M., Maxson, A., Mowson, R. & et al. 2023 Immunization Profile Study. National Association of County and City Health Officials.

9. Olusanya, O. A. et al. Sociodemographic Trends and Correlation between Parental Hesitancy towards Pediatric COVID-19 Vaccines and Routine Childhood Immunizations in the United States: 2021-2022 National Immunization Survey-Child COVID Module. Vaccines 12, 495 (2024).

10. Smith, P. J., Humiston, S. G., Parnell, T., Vannice, K. S. & Salmon, D. A. The Association Between Intentional Delay of Vaccine Administration and Timely Childhood Vaccination Coverage. Public Health Rep. 125, 534–541 (2010).

11. CDC. Measles Cases and Outbreaks. Measles (Rubeola) https://www.cdc.gov/measles/data-research/index.html (2025).

12. Eldeib, D. & Callahan, P. Why You Should Also Worry About Whooping Cough Amid Measles Outbreak. ProPublica (2025).

13. CDC. Child and Adolescent Immunization Schedule by Age. Vaccines & Immunizations https://www.cdc.gov/vaccines/hcp/imz-schedules/child-adolescent-age.html (2024).

14. Offit, P. A. & Moser, C. A. The Problem With Dr Bob’s Alternative Vaccine Schedule. Pediatrics 123, e164–e169 (2009).

15. Nguyen, K. H. et al. Trends in vaccination schedules and up-to-date status of children 19–35 months, United States, 2015–2020. Vaccine 41, 467–475 (2023).

16. Newcomer, S. R. et al. Vaccination Timeliness Among US Children Aged 0-19 Months, National Immunization Survey–Child 2011-2021. JAMA Netw. Open 7, e246440 (2024).

17. Oster, N. V. et al. A Risk Prediction Model to Identify Newborns at Risk for Missing Early Childhood Vaccinations. J. Pediatr. Infect. Dis. Soc. 10, 1080–1086 (2021).

18. Fiks, A. G. et al. Identifying Factors Predicting Immunization Delay for Children Followed in an Urban Primary Care Network Using an Electronic Health Record. Pediatrics 118, e1680–e1686 (2006).

19. Truveta. Our Approach to Data Quality. https://www.truveta.com/resources/whitepaper/truvetas-approach-to-data-quality/ (2025).

20. CDC. Catch-up Immunization Schedule for Children and Adolescents. Vaccines & Immunizations https://www.cdc.gov/vaccines/hcp/imz-schedules/child-adolescent-catch-up.html (2024).

21. HRSA. Federal Office of Rural Health Policy (FORHP) Data Files | HRSA. https://www.hrsa.gov/rural-health/about-us/what-is-rural/data-files.

22. Bright Futures/American Academy of Pediatrics. Recommendations for Preventive Pediatric Health Care. https://downloads.aap.org/AAP/PDF/periodicity_schedule.pdf?_gl=1*x2kut8*_ga*OTAxMDU2MjU5LjE3NDQ4OTYyNzU.*_ga_FD9D3XZVQQ*MTc0NDg5NjI3NS4xLjAuMTc0NDg5NjI3NS4wLjAuMA.

23. Kujawski, S. A., Yao, L., Wang, H. E., Carias, C. & Chen, Y.-T. Impact of the COVID-19 pandemic on pediatric and adolescent vaccinations and well child visits in the United States: A database analysis. Vaccine 40, 706–713 (2022).

24. Mathis, A. D. Measles Update — United States, January 1–April 17, 2025. MMWR Morb. Mortal. Wkly. Rep. 74, (2025).

25. CDC. About the National Immunization Surveys (NIS). National Immunization Surveys https://www.cdc.gov/nis/about/index.html (2025).

26. Hill, H. A. et al. Decline in Vaccination Coverage by Age 24 Months and Vaccination Inequities Among Children Born in 2020 and 2021 — National Immunization Survey-Child, United States, 2021–2023. Morb. Mortal. Wkly. Rep. 73, 844–853 (2024).

27. Dong, E., Saiyed, S., Nearchou, A., Okura, Y. & Gardner, L. M. Trends in County-Level MMR Vaccination Coverage in Children in the United States. JAMA (2025) doi:10.1001/jama.2025.8952.

28. O’Leary, S. T. et al. Strategies for Improving Vaccine Communication and Uptake. Pediatrics 153, e2023065483 (2024).

29. Limaye, R. J. et al. Communicating With Vaccine-Hesitant Parents: A Narrative Review. Acad. Pediatr. 21, S24–S29 (2021).

30. Opel, D. J. et al. The Architecture of Provider-Parent Vaccine Discussions at Health Supervision Visits. Pediatrics 132, 1037–1046 (2013).

31. Lancet, T. E. of T. Retraction—Ileal-lymphoid-nodular hyperplasia, non-specific colitis, and pervasive developmental disorder in children. The Lancet 375, 445 (2010).

32. Jain, A. et al. Autism Occurrence by MMR Vaccine Status Among US Children With Older Siblings With and Without Autism. JAMA 313, 1534–1540 (2015).

33. British Medical Journal Charges Fraud in Autism-Vaccine Paper. https://www.science.org/content/article/british-medical-journal-charges-fraud-autism-vaccine-paper.

34. Loomes, R., Hull, L. & Mandy, W. P. L. What Is the Male-to-Female Ratio in Autism Spectrum Disorder? A Systematic Review and Meta-Analysis. J. Am. Acad. Child Adolesc. Psychiatry 56, 466–474 (2017).

35. Novilla, M. L. B. et al. Why Parents Say No to Having Their Children Vaccinated against Measles: A Systematic Review of the Social Determinants of Parental Perceptions on MMR Vaccine Hesitancy. Vaccines 11, 926 (2023).

36. Zhou, E. G. et al. Parental Factors Associated With Measles–Mumps–Rubella Vaccination in US Children Younger Than 5 Years. Am. J. Public Health 115, 369–373 (2025).

37. CDC. Pertussis Surveillance and Trends. Whooping Cough (Pertussis) https://www.cdc.gov/pertussis/php/surveillance/index.html (2025).

38. Mukherjee, N. Two infants die of whooping cough in Louisiana as national case numbers climb. CNN https://www.cnn.com/2025/04/02/health/whooping-cough-pertussis-louisiana/index.html (2025).

39. Mathis, A. D. Measles — United States, January 1, 2020–March 28, 2024. MMWR Morb. Mortal. Wkly. Rep. 73, (2024).

40. CDC. About Measles. Measles (Rubeola) https://www.cdc.gov/measles/about/index.html (2025).

41. Orenstein, W., Offit, P. A., Edwards, K. M. & Plotkin, S. A. Measles. in Plotkin’s Vaccines (Elsevier, Philadelphia, 2022).

