## Supplement for "Predictors of Delayed or Absent Measles, Mumps, and Rubella Vaccination in the US, 2018–2025"

***Supplementary Methods***

Outpatient pediatric visits were defined as a visit type or class code of ‘Outpatient’ or ‘Ambulatory’, and an office visit type of: “Appointment”, “Consultation”, “Follow up”, “Hospital encounter”, “Newborn”, “Obstetrics”, “Office Visit”, “Outpatient”, “Outpatient Specialty”, “Pediatrics”, “Immunization”, “Baby”, “Transitional Care”, “Virtual Visit”, “Video Visit”, or “Urgent Care”. Excluded visit types were those classified as: “History”, “Letter”, “Patient message”, “Orders only”, “Telephone”, “Refill”, “Emergency”, “Erroneous”, where the patient could not plausibly have received an opportunity for immunization.

**Table S1.** Code Sets used to Identify Pediatric Vaccines (includes combination vaccines)

| Vaccine | CVX Codes | CPT Codes |
|---|---|---|
| **DTaP** | 20, 28, 50,102, 106, 107, 110, 115, 120, 130, 146, 170,195, 198 | 90696, 90697, 90698, 90700, 90701, 90720, 90721, 90723 |
| **HepB** | 08, 45, 51, 110, 146, 193, 198 | 90723, 90744, 90748, 90697 |
| **Hib** | 17, 22, 46, 47, 48, 49, 50, 51, 102, 120,170, 146, 198 | 90645, 90646, 90647, 90648, 90697, 90698, 90720, 90721, 90737, 90748 |
| **PCV** | 100, 133, 152, 177, 215, 216, 327 | 90669, 90670, 90671, 90677, 90732 |
| **Polio** | 10, 110, 120, 130, 146, 170, 178, 179, 195, 324 | 90696, 90697, 90698, 90712, 90713, 90723 |
| **Rotavirus** | 116, 119, 122 | 90680, 90681 |
| **MMR** | 3, 94 | 90707, 90710 |

**Table S2.** Early, On-Time, Late, and No Recorded First MMR Dose by Year of 12-21 Month Visit

| | **Year of 12-21 Month Outpatient Visit*** | | | | | | | |
|---|---|---|---|---|---|---|---|---|
| | **Overall** | **2018** | **2019** | **2020** | **2021** | **2022** | **2023** | **2024** |
| | N = 321,743 | N = 16,978 | N = 47,133 | N = 49,760 | N = 49,767 | N = 51,024 | N = 54,693 | N = 52,388 |
| **First MMR Dose** | | | | | | | | |
| Early: 6-11 months | 3,106 (1.0%) | 107 (0.6%) | 423 (0.9%) | 1,435 (2.9%) | 168 (0.3%) | 158 (0.3%) | 362 (0.7%) | 453 (0.9%) |
| On-Time: 12-15 months | 252,250 (78.4%) | 12,840 (75.6%) | 36,617 (77.7%) | 39,306 (79.0%) | 39,739 (79.9%) | 40,423 (79.2%) | 43,019 (78.7%) | 40,306 (76.9%) |
| Late: 16-23 months | 44,718 (13.9%) | 2,565 (15.1%) | 7,085 (15.0%) | 6,402 (12.9%) | 6,839 (13.7%) | 6,923 (13.6%) | 7,319 (13.4%) | 7,585 (14.5%) |
| No Vaccine by 2 Years | 21,669 (6.7%) | 1,466 (8.6%) | 3,008 (6.4%) | 2,617 (5.3%) | 3,021 (6.1%) | 3,520 (6.9%) | 3,993 (7.3%) | 4,044 (7.7%) |

*Last visit in 12–21-month window if more than one visit occurred

**Table S3.** Second Month Vaccine Timeliness by Birth Year

| | Overall | Year of Birth 2017 | 2018 | 2019 | 2020 | 2021 | 2022 | 2023 |
|---|---|---|---|---|---|---|---|---|
| | N = 321,743 | N = 48,484 | N = 44,959 | N = 51,077 | N = 50,566 | N = 54,016 | N = 54,811 | N = 17,830 |
| **First Dose Polio** | | | | | | | | |
| Early | 20,358 (6.3%) | 3,338 (6.9%) | 3,202 (7.1%) | 3,390 (6.6%) | 3,110 (6.2%) | 3,172 (5.9%) | 3,044 (5.6%) | 1,102 (6.2%) |
| On-Time | 273,086 (84.9%) | 40,486 (83.5%) | 38,058 (84.7%) | 43,959 (86.1%) | 43,102 (85.2%) | 45,894 (85.0%) | 46,637 (85.1%) | 14,950 (83.8%) |
| Late | 15,675 (4.9%) | 2,546 (5.3%) | 2,116 (4.7%) | 2,143 (4.2%) | 2,554 (5.1%) | 2,819 (5.2%) | 2,657 (4.8%) | 840 (4.7%) |
| No Dose Recorded* | 12,624 (3.9%) | 2,114 (4.4%) | 1,583 (3.5%) | 1,585 (3.1%) | 1,800 (3.6%) | 2,131 (3.9%) | 2,473 (4.5%) | 938 (5.3%) |
| **First Dose DTaP** | | | | | | | | |
| Early | 20,474 (6.4%) | 3,365 (6.9%) | 3,234 (7.2%) | 3,414 (6.7%) | 3,121 (6.2%) | 3,174 (5.9%) | 3,061 (5.6%) | 1,105 (6.2%) |
| On-Time | 274,606 (85.3%) | 40,799 (84.1%) | 38,298 (85.2%) | 44,183 (86.5%) | 43,302 (85.6%) | 46,127 (85.4%) | 46,862 (85.5%) | 15,035 (84.3%) |
| Late | 15,282 (4.7%) | 2,425 (5%) | 2,056 (4.6%) | 2,069 (4.1%) | 2,512 (5%) | 2,764 (5.1%) | 2,619 (4.8%) | 837 (4.7%) |
| No Dose Recorded* | 11,381 (3.5%) | 1,895 (3.9%) | 1,371 (3.0%) | 1,411 (2.8%) | 1,631 (3.2%) | 1,951 (3.6%) | 2,269 (4.1%) | 853 (4.8%) |
| **First Dose Hib** | | | | | | | | |
| Early | 20,172 (6.3%) | 3,318 (6.8%) | 3,178 (7.1%) | 3,356 (6.6%) | 3,084 (6.1%) | 3,134 (5.8%) | 3,016 (5.5%) | 1,086 (6.1%) |
| On-Time | 272,063 (84.6%) | 40,354 (83.2%) | 37,901 (84.3%) | 43,875 (85.9%) | 42,891 (84.8%) | 45,695 (84.6%) | 46,428 (84.7%) | 14,919 (83.7%) |
| Late | 17,452 (5.4%) | 2,803 (5.8%) | 2,394 (5.3%) | 2,361 (4.6%) | 2,862 (5.7%) | 3,122 (5.8%) | 2,992 (5.5%) | 918 (5.1%) |
| No Dose Recorded* | 12,056 (3.7%) | 2,009 (4.1%) | 1,486 (3.3%) | 1,485 (2.9%) | 1,729 (3.4%) | 2,065 (3.8%) | 2,375 (4.3%) | 907 (5.1%) |
| **First Dose PCV** | | | | | | | | |
| Early | 20,065 (6.2%) | 3,288 (6.8%) | 3,156 (7.0%) | 3,331 (6.5%) | 3,068 (6.1%) | 3,126 (5.8%) | 3,000 (5.5%) | 1,096 (6.1%) |
| On-Time | 271,003 (84.2%) | 40,147 (82.8%) | 37,756 (84.0%) | 43,690 (85.5%) | 42,803 (84.6%) | 45,585 (84.4%) | 46,181 (84.3%) | 14,841 (83.2%) |
| Late | 18,283 (5.7%) | 2,940 (6.1%) | 2,525 (5.6%) | 2,542 (5.0%) | 2,925 (5.8%) | 3,190 (5.9%) | 3,195 (5.8%) | 966 (5.4%) |
| No Dose Recorded* | 12,392 (3.9%) | 2,109 (4.3%) | 1,522 (3.4%) | 1,514 (3.0%) | 1,770 (3.5%) | 2,115 (3.9%) | 2,435 (4.4%) | 927 (5.2%) |
| **First Dose Rotavirus** | | | | | | | | |
| Early | 20,023 (6.2%) | 3,294 (6.8%) | 3,157 (7.0%) | 3,316 (6.5%) | 3,063 (6.1%) | 3,108 (5.8%) | 3,010 (5.5%) | 1,075 (6.0%) |
| On-Time | 269,450 (83.7%) | 39,717 (81.9%) | 37,474 (83.4%) | 43,366 (84.9%) | 42,540 (84.1%) | 45,435 (84.1%) | 46,114 (84.1%) | 14,804 (83.0%) |
| Late | 10,030 (3.1%) | 1,614 (3.3%) | 1,433 (3.2%) | 1,465 (2.9%) | 1,583 (3.1%) | 1,735 (3.2%) | 1,665 (3.0%) | 535 (3.0%) |
| No Dose Recorded* | 22,240 (6.9%) | 3,859 (8.0%) | 2,895 (6.4%) | 2,930 (5.7%) | 3,380 (6.7%) | 3,738 (6.9%) | 4,022 (7.3%) | 1,416 (7.9%) |

*No dose recorded by 2 years of age

**Table S4.** Fourth Month Vaccine Timeliness by Birth Year

| | | Year of Birth | | | | | | |
|---|---|---|---|---|---|---|---|---|
| | **Overall** | **2017** | **2018** | **2019** | **2020** | **2021** | **2022** | **2023** |
| | N = 321,743 | N = 48,484 | N = 44,959 | N = 51,077 | N = 50,566 | N = 54,016 | N = 54,811 | N = 17,830 |
| **Second Dose Polio** | | | | | | | | |
| Early | 7,099 (2.2%) | 1,111 (2.3%) | 1,144 (2.5%) | 1,287 (2.5%) | 1,089 (2.2%) | 1,063 (2.0%) | 1,032 (1.9%) | 373 (2.1%) |
| On-Time | 267,491 (83.1%) | 39,770 (82%) | 37,760 (84%) | 42,844 (83.9%) | 42,535 (84.1%) | 44,611 (82.6%) | 45,370 (82.8%) | 14,601 (81.9%) |
| Late | 32,229 (10%) | 5,073 (10.5%) | 4,193 (9.3%) | 5,030 (9.8%) | 4,792 (9.5%) | 5,842 (10.8%) | 5,553 (10.1%) | 1,746 (9.8%) |
| No Dose Recorded* | 14,924 (4.6%) | 2,530 (5.2%) | 1,862 (4.1%) | 1,916 (3.8%) | 2,150 (4.3%) | 2,500 (4.6%) | 2,856 (5.2%) | 1,110 (6.2%) |
| **Second Dose DTaP** | | | | | | | | |
| Early | 7,151 (2.2%) | 1,126 (2.3%) | 1,156 (2.6%) | 1,297 (2.5%) | 1,098 (2.2%) | 1,062 (2.0%) | 1,036 (1.9%) | 376 (2.1%) |
| On-Time | 268,912 (83.6%) | 40,072 (82.6%) | 37,982 (84.5%) | 43,065 (84.3%) | 42,706 (84.5%) | 44,829 (83%) | 45,593 (83.2%) | 14,665 (82.2%) |
| Late | 32,042 (10%) | 4,995 (10.3%) | 4,150 (9.2%) | 4,999 (9.8%) | 4,795 (9.5%) | 5,813 (10.8%) | 5,525 (10.1%) | 1,765 (9.9%) |
| No Dose Recorded* | 13,638 (4.2%) | 2,291 (4.7%) | 1,671 (3.7%) | 1,716 (3.4%) | 1,967 (3.9%) | 2,312 (4.3%) | 2,657 (4.8%) | 1,024 (5.7%) |
| **Second Dose Hib** | | | | | | | | |
| Early | 7,177 (2.2%) | 1,112 (2.3%) | 1,144 (2.5%) | 1,284 (2.5%) | 1,072 (2.1%) | 1,069 (2.0%) | 1,092 (2.0%) | 404 (2.3%) |
| On-Time | 266,062 (82.7%) | 39,642 (81.8%) | 37,532 (83.5%) | 42,705 (83.6%) | 42,300 (83.7%) | 44,369 (82.1%) | 44,997 (82.1%) | 14,517 (81.4%) |
| Late | 33,783 (10.5%) | 5,253 (10.8%) | 4,492 (10%) | 5,245 (10.3%) | 5,064 (10%) | 6,051 (11.2%) | 5,875 (10.7%) | 1,803 (10.1%) |
| No Dose Recorded* | 14,721 (4.6%) | 2,477 (5.1%) | 1,791 (4.0%) | 1,843 (3.6%) | 2,130 (4.2%) | 2,527 (4.7%) | 2,847 (5.2%) | 1,106 (6.2%) |
| **Second Dose PCV** | | | | | | | | |
| Early | 7,019 (2.2%) | 1,103 (2.3%) | 1,131 (2.5%) | 1,272 (2.5%) | 1,070 (2.1%) | 1,049 (1.9%) | 1,023 (1.9%) | 371 (2.1%) |
| On-Time | 264,919 (82.3%) | 39,359 (81.2%) | 37,379 (83.1%) | 42,492 (83.2%) | 42,202 (83.5%) | 44,211 (81.8%) | 44,800 (81.7%) | 14,476 (81.2%) |
| Late | 34,801 (10.8%) | 5,497 (11.3%) | 4,613 (10.3%) | 5,450 (10.7%) | 5,146 (10.2%) | 6,208 (11.5%) | 6,105 (11.1%) | 1,782 (10%) |
| No Dose Recorded* | 15,004 (4.7%) | 2,525 (5.2%) | 1,836 (4.1%) | 1,863 (3.6%) | 2,148 (4.2%) | 2,548 (4.7%) | 2,883 (5.3%) | 1,201 (6.7%) |
| **Second Dose Rotavirus** | | | | | | | | |
| Early | 7,162 (2.2%) | 1,085 (2.2%) | 1,137 (2.5%) | 1,291 (2.5%) | 1,079 (2.1%) | 1,090 (2.0%) | 1,084 (2.0%) | 396 (2.2%) |
| On-Time | 263,515 (81.9%) | 38,957 (80.4%) | 37,117 (82.6%) | 42,204 (82.6%) | 41,938 (82.9%) | 44,119 (81.7%) | 44,784 (81.7%) | 14,396 (80.7%) |
| Late | 22,219 (6.9%) | 3,366 (6.9%) | 2,909 (6.5%) | 3,617 (7.1%) | 3,212 (6.4%) | 3,957 (7.3%) | 3,919 (7.2%) | 1,239 (6.9%) |
| No Dose Recorded* | 28,847 (9.0%) | 5,076 (10.5%) | 3,796 (8.4%) | 3,965 (7.8%) | 4,337 (8.6%) | 4,850 (9.0%) | 5,024 (9.2%) | 1,799 (10.1%) |

*No dose recorded by 2 years of age

**Table S5.** Output for Model of No MMR Vaccination by 2 Years vs. Any MMR Vaccination

| | Before COVID-19 Pandemic | | After COVID-19 Pandemic | |
|---|---|---|---|---|
| ***Covariate*** | ***Estimate*** | ***Confidence Interval*** | ***Estimate*** | ***Confidence Interval*** |
| **AAP Well Child Visit Schedule** | | | | |
| Not Followed | REF | | REF | |
| Followed | 0.37 | (0.33, 0.40) | 0.37 | (0.36, 0.39) |
| **Year of 12-21 Month Visit** | | | | |
| 2018 | REF | | | |
| 2019 | 0.71 | (0.66, 0.77) | -- | -- |
| 2020 | 0.74 | (0.65, 0.83) | REF | |
| 2021 | -- | -- | 0.97 | (0.89, 1.06) |
| 2022 | -- | -- | 1.18 | (1.08, 1.29) |
| 2023 | -- | -- | 1.19 | (1.09, 1.30) |
| 2024 | -- | -- | 1.35 | (1.23, 1.47) |
| **Second Month Vaccine Timing** | | | | |
| On-Time | REF | | REF | |
| Late | 7.73 | (7.03, 8.51) | 6.96 | (6.60, 7.34) |
| **Fourth Month Vaccine Timing** | | | | |
| On-Time | REF | | REF | |
| Late | 6.56 | (5.94, 7.24) | 6.16 | (5.84, 6.50) |
| **Rural/Urban Residence** | | | | |
| Urban | REF | | REF | |
| Rural | 1.18 | (1.07, 1.30) | 1.09 | (1.03, 1.15) |
| Unknown | 1.10 | (0.75, 1.61) | 1.45 | (1.18, 1.79) |
| **Sex** | | | | |
| Female | REF | | REF | |
| Male | 1.11 | (1.04, 1.20) | 1.12 | (1.07, 1.16) |
| **Race** | | | | |
| Black | REF | | REF | |
| Asian | 0.87 | (0.70, 1.08) | 0.81 | (0.71, 0.91) |
| White | 1.40 | (1.23, 1.59) | 1.48 | (1.39, 1.59) |
| Other | 1.15 | (0.96, 1.38) | 1.13 | (1.02, 1.25) |
| Unknown | 1.42 | (1.19, 1.71) | 1.22 | (1.11, 1.33) |
| **Ethnicity** | | | | |
| Hispanic or Latino | REF | | REF | |
| Not Hispanic or Latino | 1.33 | (1.16, 1.52) | 1.51 | (1.41, 1.61) |
| Other | 0.78 | (0.53, 1.16) | 1.07 | (0.91, 1.27) |
| Unknown | 1.26 | (1.06, 1.49) | 1.43 | (1.31, 1.57) |

**Table S6.** Output for Model of Late MMR Vaccination vs. Timely MMR Vaccination

| | Before COVID-19 Pandemic | | After COVID-19 Pandemic | |
|---|---|---|---|---|
| ***Covariate*** | ***Estimate*** | ***Confidence Interval*** | ***Estimate*** | ***Confidence Interval*** |
| **AAP Well Child Visit Schedule** | | | | |
| Not Followed | REF | | REF | |
| Followed | 0.74 | (0.71, 0.78) | 0.61 | (0.59, 0.63) |
| **Year of 12-21 Month Visit** | | | | |
| 2018 | REF | | -- | -- |
| 2019 | 0.97 | (0.92, 1.02) | -- | -- |
| 2020 | 0.66 | (0.61, 0.72) | REF | |
| 2021 | -- | -- | 0.87 | (0.82, 0.92) |
| 2022 | -- | -- | 0.89 | (0.84, 0.95) |
| 2023 | -- | -- | 0.88 | (0.83, 0.94) |
| 2024 | -- | -- | 1.00 | (0.95, 1.06) |
| **Second Month Vaccine Timing** | | | | |
| On-Time | REF | | REF | |
| Late | 1.51 | (1.35, 1.68) | 1.47 | (1.37, 1.57) |
| **Fourth Month Vaccine Timing** | | | | |
| On-Time | REF | | REF | |
| Late | 2.46 | (2.27, 2.66) | 2.55 | (2.43, 2.67) |
| **Rural/Urban Residence** | | | | |
| Urban | REF | | REF | |
| Rural | 0.87 | (0.82, 0.93) | 0.69 | (0.66, 0.71) |
| Unknown | 1.29 | (0.98, 1.71) | 1.06 | (0.89, 1.26) |
| **Sex** | | | | |
| Female | REF | | REF | |
| Male | 1.04 | (0.99, 1.09) | 1.01 | (0.99, 1.04) |
| **Race** | | | | |
| Black | REF | | REF | |
| Asian | 0.53 | (0.45, 0.61) | 0.47 | (0.43, 0.51) |
| White | 0.98 | (0.90, 1.06) | 0.87 | (0.83, 0.92) |
| Other | 0.79 | (0.70, 0.90) | 0.77 | (0.72, 0.83) |
| Unknown | 0.82 | (0.73, 0.93) | 0.80 | (0.75, 0.85) |
| **Ethnicity** | | | | |
| Hispanic or Latino | REF | | REF | |
| Not Hispanic or Latino | 1.09 | (1.00, 1.19) | 1.25 | (1.19, 1.31) |
| Other | 1.10 | (0.85, 1.43) | 1.18 | (1.03, 1.36) |
| Unknown | 1.15 | (1.03, 1.29) | 1.42 | (1.33, 1.51) |
